# Leveraging Large Language Models to Direct Automated PET/CT Tumour Segmentation in Retrospective Data: an Agentic Framework Method

**DOI:** 10.1101/2025.11.27.25339233

**Authors:** Lara M. Bonney, Stuart L. Brown, Kevin M. Bradley, Andrew B. Hassan, Daniel R. McGowan

**Affiliations:** Sir William Dunn School of Pathology, University of Oxford; Department of Medical Physics and Clinical Engineering, Oxford University Hospitals NHS Foundation Trust; Wales Research and Diagnostic Positron Emission Tomography Imaging Centre (PETIC), University Hospital of Wales; Department of Oncology, University of Oxford

## Abstract

Automated medical image segmentation using deep learning requires large labelled datasets, presenting barriers for rare cancers like sarcoma. We developed an agentic framework integrating LLM analysis of radiologist reports with nnUNet segmentation for ^18^F-FDG PET/CT imaging data (N=60, 134 studies), aiming to improve automated tumour segmentation for retrospective data. LLM interpretation was optimised in an expanded report dataset (N=91, 226 studies). The framework aimed to first screen for disease presence, then tested localising tumours to body regions mapped via automated organ segmentation, enabling targeted image cropping before segmentation. A baseline nnUNet was trained for comparison and achieved a mean Dice of 0.49. The disease classification framework attained mean Dice 0.62, correctly identifying 9/11 disease-free cases versus 6/11 for baseline. This approach demonstrates potential for leveraging routinely collected clinical text data to enhance medical imaging research in rare diseases.

## 1 Introduction

Retrospective healthcare data contains a wealth of information that can be mined for research, however, data cleaning and labelling is resource intensive and remains a significant barrier to use. In the context of imaging, data labelling typically refers to manual segmentation of volumes of interest. Deep learning, and in particular the UNet architecture implemented through nnUNet, has fast become the baseline for automated medical image segmentation [1]. However, undifferentiated tumour segmentation is a challenging task, with high heterogeneity in the data at both an inter- and intra-patient level [2]. To counteract this either large labelled training datasets or user input is required to limit the algorithm to a smaller region of interest for optimal results. For large datasets this level of manual input, although less than for completely manual segmentation, remains resource intensive and potentially prohibitive for large scale applications.

This challenge is particularly acute for rare cancers, which collectively represent a substantial disease burden yet due to low incidence rates receive disproportionately limited research investment. The data focus in the era of AI is compounding this disparity in two parts. Firstly, for rare cancers, gathering large enough datasets to represent the heterogeneity in patient presentation can present additional challenges which is a barrier to research in such areas [3]. Secondly, where development primarily focuses on benchmark datasets favouring common conditions with abundant training examples, rare diseases are systematically under-represented in algorithmic advancement [4]. Sarcoma exemplifies this challenge, encompassing over 70 different histological subtypes with heterogeneous biological behaviours that can occur anywhere in the body, yet accounting for only 2% of adult malignancies [5], [6]. It is a disease that has high potential for benefit from precision medicine, but precision medicine requires a multi modal approach with a sufficient evidence base that is challenging to build. Imaging data is a key piece of information in this process, providing vital information on treatment response, while demonstrating additional predictive properties in the literature [7]. A multi-omic approach in sarcoma, including imaging data, is particularly relevant due to the size and varied location of tumours. However, utilising this data at scale is inhibited by the resource intensive segmentation required.

Most automated medical image segmentation research focuses on prospective image-only (“vision”) segmentation, which offers the greatest potential benefit for clinical radiology workflows. However, mining retrospective healthcare data presents substantial utility for research and service development. In this context the accompanying radiologist report is also available. This direct interpretation of the image by a highly trained specialist is temporally linked to the imaging data and provides clinical context to the imaging that has high potential benefit in data driven approaches. On the other hand, radiology reports are unstructured free-text data which require additional interpretation/processing for use. Numerous studies have shown that large language models (LLMs) can be used with a high degree of success to extract key features from radiology reports [8], [9]. This capability can be utilised in multi-modal approaches such as visual language models (VLMs) which have shown significant performance improvements in recent radiology focused publications [10], [11], [12], [13]. However, VLMs typically require large-scale datasets for optimal performance, limiting the applicability to specialised tasks or rare pathologies where such datasets are unavailable.

An alternative approach lies in agentic AI systems, which engineer frameworks around LLMs and other AI models to access accurate knowledge, apply reasoning and take actions [14]. These systems dynamically select appropriate analytical approaches based on available information, making them particularly suited to scenarios with limited training data. Several studies have begun to explore the use of agentic frameworks in radiology with high levels of success [15], [16].

This work explores an agentic framework method for tumour segmentation in a retrospective PET/CT sarcoma dataset where both image data and the accompanying text based radiologist report is available. The work tests the hypothesis that LLM interpretation of radiologist reports can improve automated tumour segmentation performance, whilst exploring the implementation and challenges of developing such a framework in a small dataset, with intended application in rare disease research.

## 2 Materials and Methods

### 2.1 Dataset

The data used in this work is from the Oxford Precision Oncology for Sarcoma trial (OxPOS) [22/SC/0054]. This is a prospective observational real world data study with access to patients retrospective clinical care data. For inclusion in the study patients must be 18 or older, have a high-grade sarcoma (Grade 2 or 3) on either a biopsy or surgical sample confirmed by the Oxford sarcoma MDT, have a life expectancy greater than 12 weeks from the point of diagnosis, and be able to undergo all clinical treatments and follow-up delivered by the Oxford Sarcoma supra-regional MDT. The imaging data used in this work contains all ^18^F-FDG PET/CT studies from the first 60 patients recruited to OXPOS. PET/CT imaging consists of two separate modalities, with the PET image a 3D map of the radiolabelled tracer in the body (in this instance ^18^F-FDG) and the CT a 3D anatomical image. The imaging dataset contained 132 PET/CT labelled sarcoma studies. The standard imaging extent is skull base to upper thigh. For patients with lower limb tumour volumes the imaging extent is either extended or a separate standalone limb image is acquired. The dataset included 36 studies with standalone lower limb imaging, giving a total of 168 PET/CT image pairs (336 images). This dataset comprises staging, treatment response and disease monitoring scans. Labelling was performed by an experienced dual qualified Nuclear Medicine radiologist with 23 years experience in the field (KB). In all studies the existence of disease and location of disease agreed with the radiologist report at the time of the study. The data pre-processing steps completed are summarised in Supplementary Figure 1.

The dataset was split into train and test sets with an 80:20 split. The split was balanced on total number of studies, number of PET/CT studies per patient, disease present in image, and the number of limb images. The split is shown in Table 1.

**Table 1.**
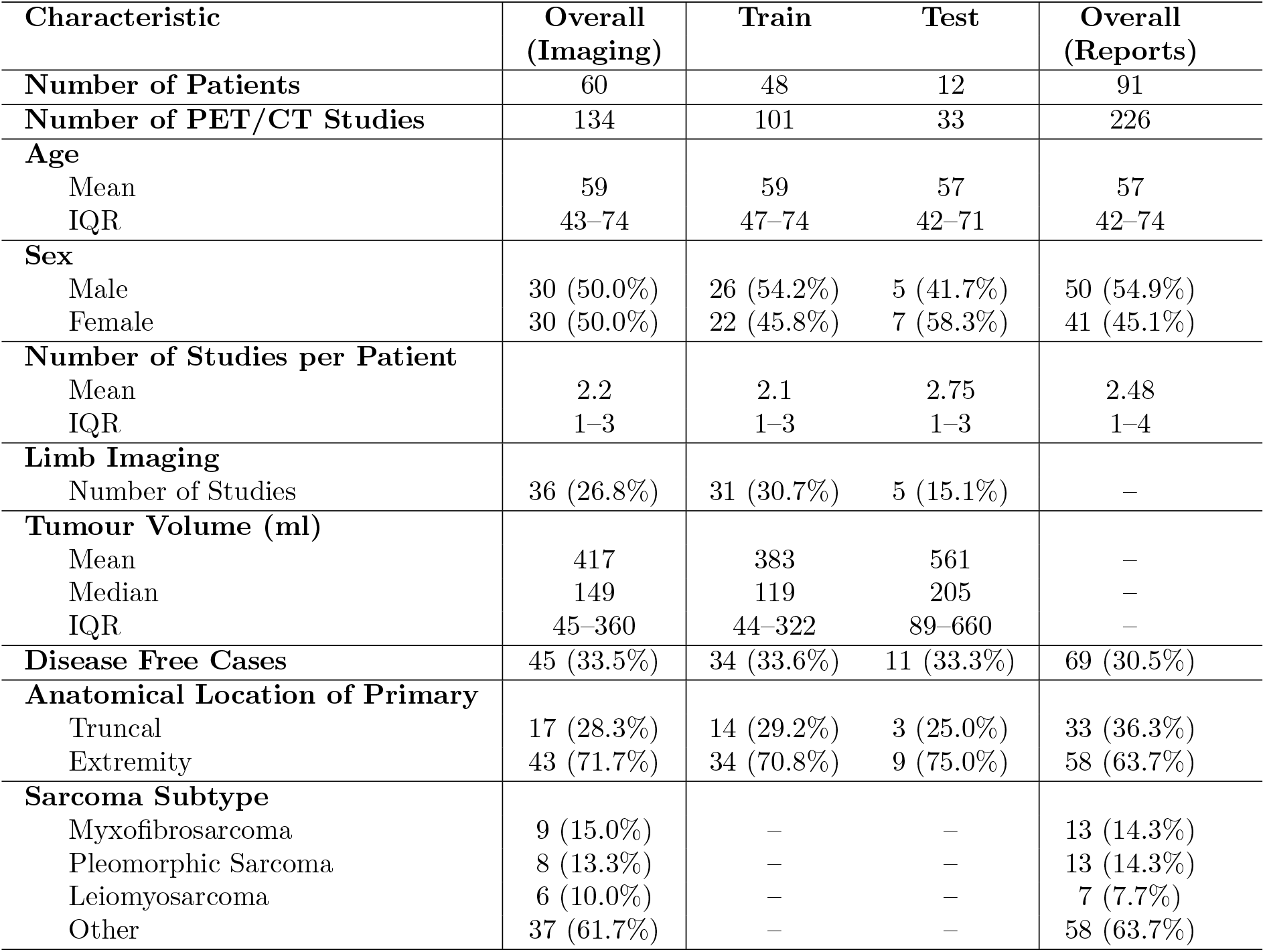
Population characteristics between the test and train data for segmentation algorithm development and radiology report interpretation. For the first 60 patients recruited to the study all PET/CT images were segmented by an experience nuclear medicine radiologist this formed the imaging dataset for this work. For optimisation of radiology report interpretation, all patients recruited at the date analysis began were included. In the imaging dataset a random selector was run to split patients into an 80:20 split balanced on number of studies per patient, limb imaging, disease free cases, and anatomical location. Groups of five or less have not been specified for the specific diagnosis to retain anonymisation.

An expanded dataset with the first 91 patients recruited to OXPOS was used for the LLM testing. This dataset included the radiology reports for the above images, and a further 93 reports with an additional 31 patients (a number of the further reports are for studies acquired since the original data collection in the initial 60 patients), giving a total of 226 radiology reports from the 91 patients. The reports were interpreted against two questions. This interpretation was validated against the segmentation results for those cases with ground truth segmentation. No language model training or fine-tuning was undertaken and as such the whole dataset was used for testing the LLM task specific performance.

#### Performance Metrics

To evaluate the overall segmentation performance, the following metrics were assessed in the test dataset. Dice score, as defined in Equation 1 for the ground truth volume, G_t_, and predicted volume, P. Where lower limb imaging was completed separately the Dice score was calculated over the total imaging volume for that study to give a single data point. The total false positive and false negative volumes. Finally, the proportion of negative cases correctly predicted as negative - although this is reflected in the other metrics, it has a particularly high impact and is useful for understanding results.

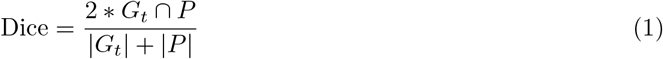

The metrics used for optimising decision points in the framework are detailed in the relevant sections.

#### Baseline Model

A baseline nnUNet model was trained using best strategies established in autoPET. The data was labelled with five organ classes and tumour label data. The nnUNet trainer was customised to oversample the tumour class at a rate of 33% and training was run for 1500 epochs [17]. Notably, the distribution was bimodal, with the model either performing well or very poorly in most cases. With the very poor performance cases primarily contributed to by a small number of very high false negative or false positive volumes. The test dataset contains 11 cases without disease present, these cases can significantly skew the Dice distribution as they either measure a 0 or a 1. While this is a limitation of Dice it is also useful for tracking performance as it has high clinical significance. The baseline model correctly predicted 6 of the 11 cases (54%). The baseline model incorrectly predicted 2 of the 22 positive cases as containing no disease, also returning a Dice coefficient of 0.

A model was also trained using the same parameters on cropped image data for use in the localisation workflow. This model performed better when only the tumour class was labelled, this is to be expected as the cropping removes the context provided by organ labels.

#### Proposed Framework

The primary problem posed by the use of a limited dataset for a rare disease with heterogeneous presentation was a high false positive rate. The proposed agentic framework looks to counter high false positive volumes through two levels of LLM interpretation. First, the framework looks to screen out studies which do not contain any volume(s) for segmentation, then for cases which contain volume(s) for segmentation it looks to define the region of the body where these volume(s) are. These regions can then be mapped to the images using automated organ segmentation methods, with the images cropped to this region prior to the application of the segmentation model. This process is summarised in Figure 1.

**Figure 1.**
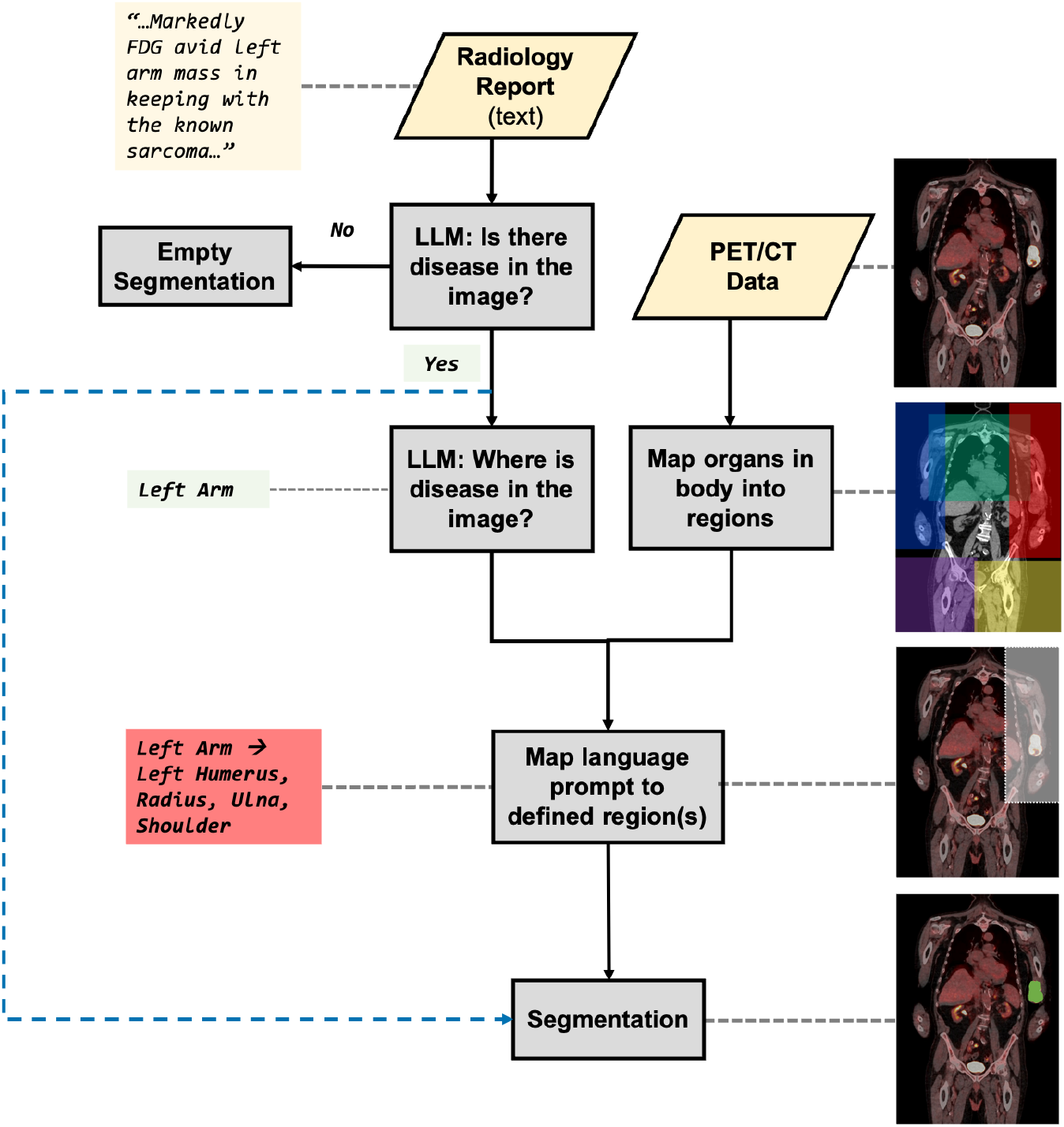
Agentic Report Informed Segmentation Framework: Overview flowchart of the framework with example data input. This work tests two different levels of report interpretation to the segmentation process. Results are reported with and without the localisation task (denoted by the dashed arrow).

The following sections outline the set-up of this framework and how the interpretation of the two LLM tasks was optimised. It should be noted that there are a large number of parameters that could be optimised in this process, and it is not feasible to test all parameters. The work has focused on the areas identified by the authors as offering the maximum return.

#### Large Language Model Radiology Report Interpretation

To utilise the radiology reports in this dataset for an automated segmentation framework requires a high amount of data processing due to the unstructured, free-text nature of reports. In addition reports are written in different styles by different clinicians and are often answering a specific question different to the question wanted for the purpose of research. In this dataset reports end with a short conclusion/interpretation section, therefore, the LLM testing investigated including the full report or only the conclusion section in the prompt (text vs. conclusion).

The framework requires the LLM output to be in a standardised format for ongoing interpretation, thus favouring an instruction-tuned LLM. Instruction tuned LLMs are fine-tuned on a labelled dataset of prompts and corresponding responses [8]. This improves model performance not only on the specific tasks presented in the training dataset but also on following instructions in general. For the purpose of this work ≤8B open source models were preferred due to compute availability and data security. Three standard and three medically fine-tuned models were tested in this work.

1. Meta Llama 3.1 8B Instruct [18]
2. Mistral 7B Instruct version 0.3 [19]
3. Qwen 2.5 7B Instruct [20]
4. OpenBioLLM 8B - fine tuned on Meta Llama 3 8B Instruct [21]
5. BioMistral 7B - fine tuned on Mistral 7B Instruct [22]
6. Qwen 2.5 7B Instruct UMLS - fine tuned on Qwen 2.5 7B Instruct [23]

Two questions were asked of each report, and were formulated into a prompt that instructed the model to only respond from the pre-defined list of answers. The list of answers was chosen for both tasks to be in plain English to maximise the chance of the model recognising and using the output. The first task is relevant to a wide range of clinical imaging indications while the second task has high clinical relevance to sarcoma, a disease which occurs throughout the body in a wide array of anatomical locations.

1. **Disease Classification:** Is there evidence of FDG avid disease in the PET/CT study? (Binary)
2. **Localisation:** Where is the tumour, recurrence or metastatic disease in the study? (Pre-defined list of anatomical regions, and option of “No disease”)

The LLM responses were cleaned against the accepted answer list, a strict cleaning was applied. Aiming to reduce the impact of hallucinations the “confidence” of the LLM in the answer was tested. Each question was asked *N* = 12 times with slightly altered wording, producing a set of predictions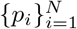. The exact questions asked are shown in Supplementary Material Tables 3 and 4. The model did not have any memory of the previous prompt or response. The final answer was taken as the modal response, 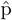, with confidence score, c(x), defined as:

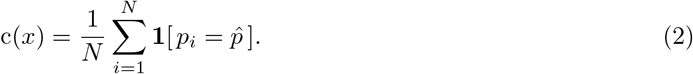

It is hypothesised that setting a threshold confidence score below which the LLM response is discarded and the baseline assumption taken will improve the sensitivity/specificity of the overall framework. The baseline assumption is that there is FDG avid disease present in the image, and the disease could be anywhere in the body. This encodes that if the model has high confidence then it will predict and use the result, but if the model has low confidence then it will fall back to the model default. For example, in the event that the confidence threshold is not met for either question the image will be fed to the nnUNet baseline model without any adaptation relying on the predictive capability of the model. Figure 2a demonstrates this and the alternative data flows where the confidence threshold is met.

**Figure 2.**
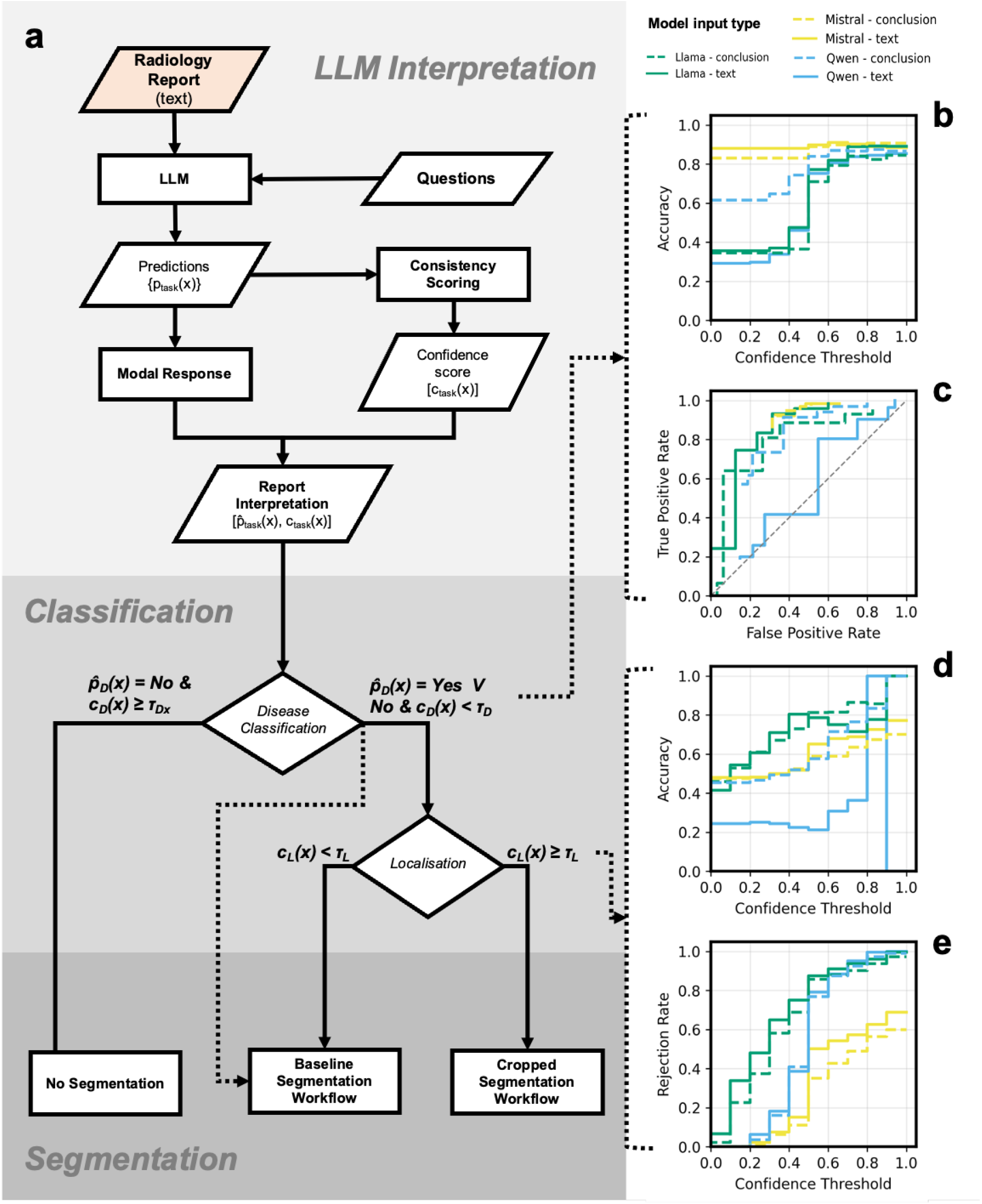
Optimisation of Large Language Model Input to Segmentation Workflow. **a:** Overview of proposed framework for LLM response interpretation and implementation. Baseline segmentation workflow refers to the baseline nnUNet model being applied to the full image, while the cropped segmentation model process is shown in further detail in Figure 3. The results were also calculated for only including the disease classification task in the overall pipeline, as shown by the dashed line. **b:** Accuracy of the overall classification against confidence threshold. **c:** Receiver Operator Characteristic Curve for the overall process, the plot does not pass through (0,0) because the decision variable is not continuous. **d:** Accuracy of the localisation task as a function of confidence threshold. **e:** Rejection rate of the localisation task as a function of confidence threshold.

To select the optimal confidence threshold for the binary outcome data the sensitivity and specificity of each confidence threshold, *τ*, was assessed. For each report, x, the predicted value, p(x), with confidence, c(x), were compared to the ground truth value g_t_(x) with true positives (TP), true negatives (TN), false positives (FP) and false negatives (FN) defined as per Equation 3. The definition of the confusion matrix is comprehensive, however, because the decision threshold is not continuous the receiver operator characteristic (ROC) curves do not necessarily extend to the extrema of true and false positive rates in all cases.

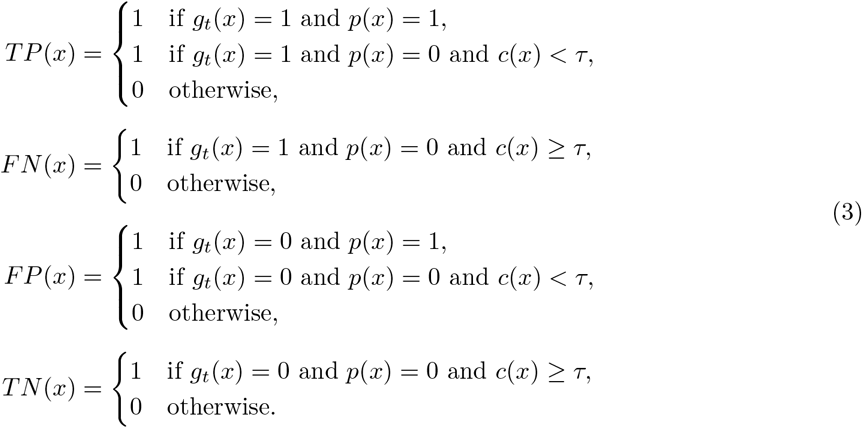

The performance of the localisation task was assessed using the accuracy and rejection rate due to the non-binary output. The accuracy was defined as the proportion of cases with confidence exceeding the threshold that are inclusive of the correct response as per Equation 4. This definition does not penalise for the model response predicting additional regions, clinically this is relevant as it reduces the likelihood of the overall framework missing a relevant region of the body.

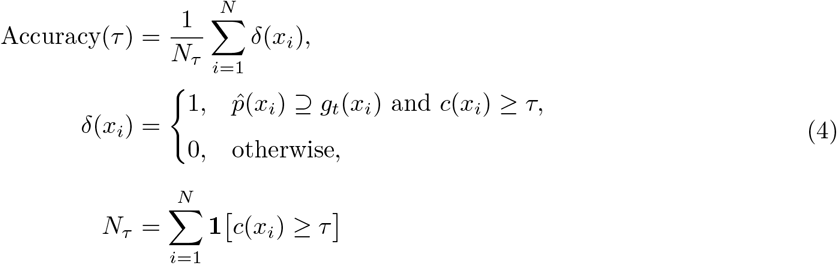

#### Linking Text Based Localisation to Anatomy

Since sarcoma is a highly heterogeneous disease in presentation, it is hypothesized that using the LLM to localise the segmentation output could improve results. Image segmentation work in literature has shown marked performance improvements where the full image is cropped prior to presentation to the model, narrowing the field of view and centering on the region of interest. In large retrospective datasets this can require a level of user input that remains prohibitively time consuming for research. Here, we aimed to crop the image based on the radiology report interpretation mapped to organs from an automated organ segmentation model, Total Segmentator [24]. The regions of the body and organ mapping from Total Segmentator included in the regions are listed in Supplementary Material Table 2. The proposed workflow is shown in panel A of Figure 3. A buffer zone was added to the organ cropping bounding box to increase the likelihood that the total tumour region would be included. The buffer used was varied dependent on the position of the anatomical region in question. To assess the suitability of the crop regions the containment fraction was defined as the fraction of the predicted volume, P(x), contained within the body region volume, R(x). Where the LLM assessment returned multiple body regions for assessment, these were cropped and combined to form a total R(x) for assessing the containment fraction. In the framework individual regions are fed to the segmentation model with the outputs combined at the end.

**Figure 3.**
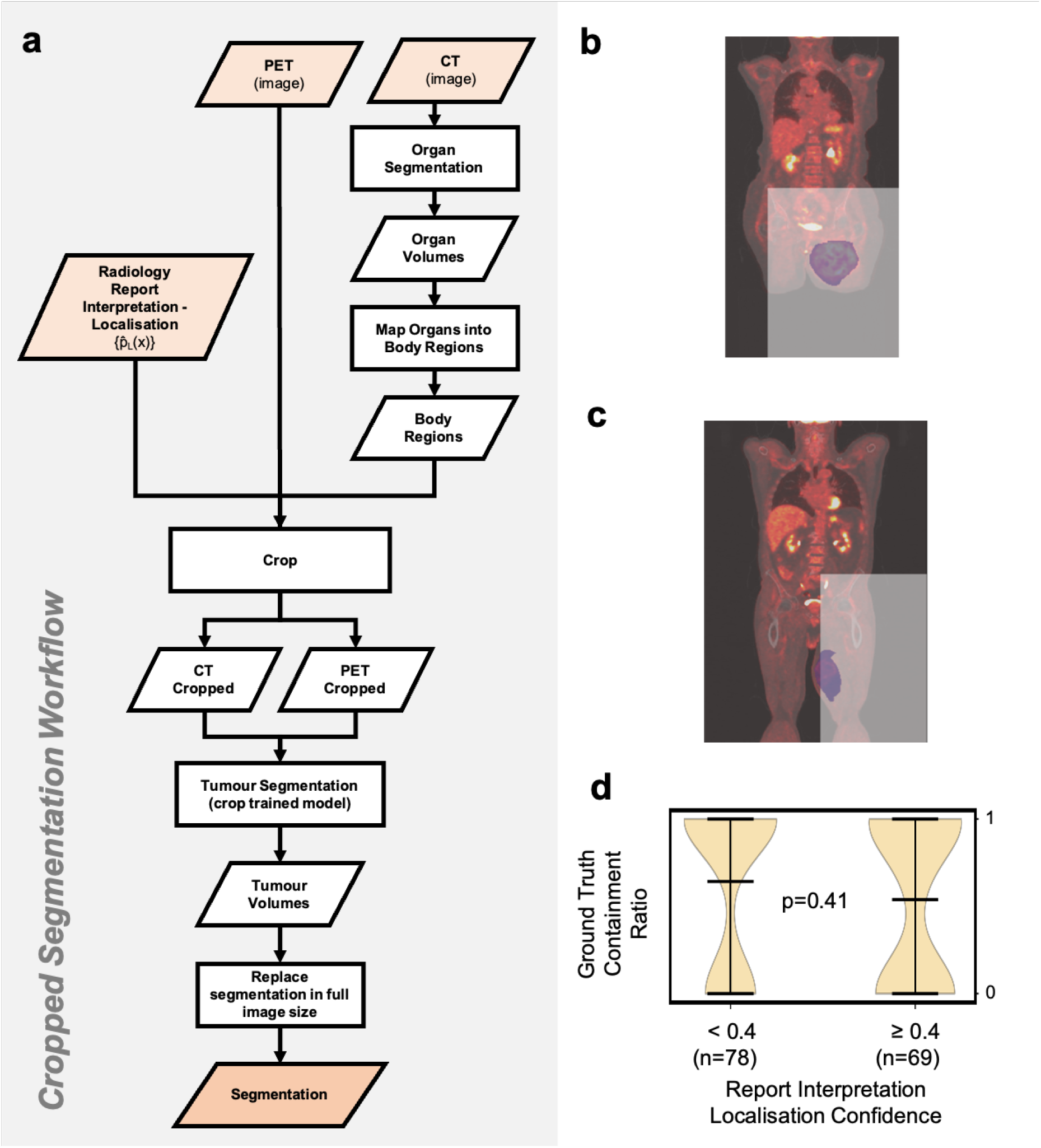
Optimisation of Localisation Framework. **a:** The flowchart shows the method used to incorporate LLM interpretation of localisation into the framework. **b:** Left thigh pleomorphic sarcoma, fused PET/CT image with tumour segmentation shown in purple and localised region shown in white. Accurate LLM prediction of “Left Leg” with ground truth containment fraction of 1.0 in the left leg. The localised region in this case is oversized and extends into the right leg. **c:** Left thigh liposarcoma, fused PET/CT image with tumour segmentation shown in purple and localised region shown in white. Example case of accurate LLM prediction “Left Leg” with ground truth containment fraction of 0.90 in the left leg. **d:** Violin plot of the distribution of containment ratio of the ground truth volume split of the LLM confidence score. There was no significant difference between the two groups (p=0.41).

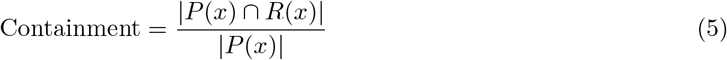

#### Overall Framework Performance

The significance of the difference between performance of the baseline model to the disease classification only framework and the overall framework including text based localisation was tested using a Wilcoxon signed rank test (*α*=0.05) for each performance metric over the distribution of test cases.

## 3 Results

### Large Language Model Radiology Report Interpretation

The primary aim of the disease classification task is to detect cases without disease, such that these images are not passed to the nnUNet model with the overall goal of decreasing false positive cases. Therefore, a high specificity is important. However, the primary objective for optimising this task is a high sensitivity, such that patients with disease are not misclassified resulting in false negative cases. All models demonstrated a trend towards increased accuracy with higher confidence threshold. The Mistral performance was also the most stable to confidence with a narrow range of sensitivity and specificity values achieved. The optimal model performance was deemed to be Mistral conclusion only with a confidence threshold of 0.6, achieving a sensitivity of 0.93, specificity of 0.69, and overall accuracy of 0.89, Figure 2b,c. Across all the models there was no clear trend between including the full radiology text or conclusion only. No consistent difference was observed between medically fine-tuned and general models, for simplicity results are displayed for general models only, with medically fine-tuned results available in Supplementary Figure 2.

For the task of localisation the accuracy rate was lower overall, which is as expected due to the higher complexity of the task. The confidence values measured were also overall lower with high rejection rates observed as the confidence threshold was raised, particularly for the Llama and Qwen models. To avoid the introduction of false negative segmentation the task requires a high accuracy rate, with a high reject rate acceptable as the baseline model can be used in these cases. The best performing model was deemed to be Llama using the full radiology report at a confidence threshold of 0.4, this achieved an accuracy of 0.81 for a rejection rate of 0.76.

#### Linking Text Based Localisation to Anatomy

Combining the LLM localisation task response and automated organ segmentation defined body regions to define a crop region was found to be moderately accurate. Achieving an mean ground truth containment fraction of 0.57 across the 147 images tested, images where the LLM predicted no disease could not be included in this test. Notably, there was no significant difference (p=0.41) in the cohorts between confident and less confident LLM responses, with a higher mean ground truth containment fraction recorded for low confidence answers (0.65 vs. 0.54 respectively). The distribution of containment fraction was bimodal for both high and low confidence answers, Figure 3c, and can be linked to the incorrect LLM responses. This is likely to induce false negatives when deployed into the framework.

#### Overall Framework Performance

The framework with disease classification achieved a mean Dice of 0.62 [median: 0.89] with a negatively skewed distribution, correctly identifying 9 out of 11 disease free cases in the test dataset. In the image only nnUNet models the best mean Dice achieved was 0.49 [median: 0.65], which was considerably biased by a high number of false positive disease free cases (negative case prediction rate 6 out of 11, Dice=0.0). The difference did not reach statistical significance (Wilcoxon signed-rank test: W=3.0, p=0.053), however, the LLM disease classification method showed a small positive effect compared to baseline (Cohen’s d=0.30), with mean Dice scores improving from 0.49 to 0.62. Using the framework with localisation worsened performance, achieving a mean Dice of 0.56 [median: 0.73]. This is an improvement on the baseline model, but a worsened performance as compared to the disease classification only model. This difference arises from an increase in false negative volumes in the cases where localisation is used. The nnUNet models which were trained including organ labels (label set 2) consistently outperformed the models which were trained with only tumour label data (label set 1) when viewing the whole image, while for cropped data using only the tumour class improved performance.

## 4 Discussion

The improvement in overall segmentation performance demonstrates the potential of agentic frameworks in the context of medical image segmentation for small retrospective datasets. Although the result does not quite reach significance (p=0.053), the distribution shift is consistent (W=3.0) with a 25% improvement in mean Dice score. Comparison with image only segmentation tasks is somewhat void as additional data is presented to the framework as compared to a vision only model. However, given the size of the dataset and initial challenges, raising the mean Dice to 0.62 is highly encouraging. This value is considerably closer to the best overall Dice achieved in recent iterations of challenges such as autoPET (0.79, 2023) [2]. The most appropriate benchmark for an adequate Dice is that achieved when comparing clinicians. There are limited studies in literature of the Dice achieved when comparing different clinicians. Work by Wong et al. found a Dice of 0.72 between clinicians for pancreatic cancer CT [25]. Pancreatic tumours are notoriously challenging to segment. However, notably these tumours also cover a significantly narrower anatomical region than sarcoma, and the clinicians had the added assistance of feedback prior to final evaluation where volumes were outlying.

The results of this work have focused on Dice, which has significant limitations. In particular, the skewing of distributions from 0 and 1 values. However, in this context it was felt those cases where a Dice of 0 or 1 was measured had high clinical significance and therefore the focus on Dice was appropriate.

The overall LLM accuracy enabled improvement in segmentation performance observed. Notably, all models displayed improved accuracy with increasing confidence, which is notable as there are conflicting reports in literature as to whether this should be the case and it is not clear whether the repeatability (confidence) of LLM response is indicative of overall accuracy [26], [27]. While it may seem a surprising result that the medically fine tuned models did not outperform the general models, this has been previously observed in literature for unseen data outside of benchmarking datasets and underscores the importance of user specific testing [28].

The use of an LLM to interpret the reports is, in part, required due to the unstructured nature of the radiology reports. Proforma reporting is now common place for other oncological indications and imaging modalities [29], [30], [31]. However, in rare diseases, and a disease as heterogeneous as sarcoma, implementing proforma/structured reporting presents a number of challenges. While the LLM accuracy was high overall, a semi-structured reporting approach could enable easier post-structuring of reports for research and service improvement which could negate the requirement for using LLMs with the risk of hallucinations or mistakes. The LLM interpretation is also required as when using retrospective clinical care data for research the research question often differs significantly from the clinical question the report was written to answer. Structured data provided by proforma reporting would not necessarily solve this. Radiologist report disease classification performance was overall good and could be improved when considering the “confidence” of the model response (Figure 2). PET/CT is a complex imaging modality, with long associated radiologist reports. While the reports have an overall structure with a short conclusion section at the end, the findings of the conclusion section are inherently dependent on, and refer to, the clinical indication/referral for which the patient was sent for the scan. PET/CT is primarily used for staging, treatment response, and ongoing monitoring as opposed to early stage diagnostic imaging, and as such the reports tend to have a less specific focus on whether there is disease in the image as a whole - the question that was being asked in this task. Importantly, in this instance, the referral data was not available and could not be provided to the model, although this could be an area for potential improvement. Many VLM models for segmentation published in medical imaging are centred on publicly available datasets where the accompanying text is structured data relevant to the segmentation approach. Using a dataset that isn’t publicly available also ensures that the data will not have featured in model training or fine-tuning.

The LLM disease localisation task performance was considerably poorer and including it in the framework worsened overall segmentation performance. This worsened performance can be primarily attributed to the lower LLM task accuracy. This task is complex and the mean confidence of the task was considerably lower than for disease classification (0.35 vs 0.91). The dataset includes patients with metastatic disease, including studies with extensive disease throughout the body and translating this to localised regions is particularly challenging. The worsened performance can also be attributed to the uncertainty in organ to region mapping and edge effects. High accuracy was observed for the organ segmentation, however, the approach taken to generate cuboid regions from the outer bounds of the relevant organ segmentations experienced some inaccuracies at edges. An example of this is shown in Figure 3b,c for two patients both with Left Leg tumours. In panel b the angle of the patients femur results in an oversized region, while in panel c the angle of the patients femur results in an undersized region. This performance could be improved by incorporating body outline segmentation or use of an atlas method. However, the LLM task for localisation required a high rejection rate to achieve sufficient accuracy, this meant that from the test set the cropped segmentation workflow was only applied to 8 studies. Notably, the cropped model performs similarly on Dice to the baseline model, shown in orange in Figure 4. For this evaluation it was deployed over all regions in the body for every case. This suggests that the worsened performance arises from the LLM task performance as opposed to the region cropping or localisation although small differences are expected here.

**Figure 4.**
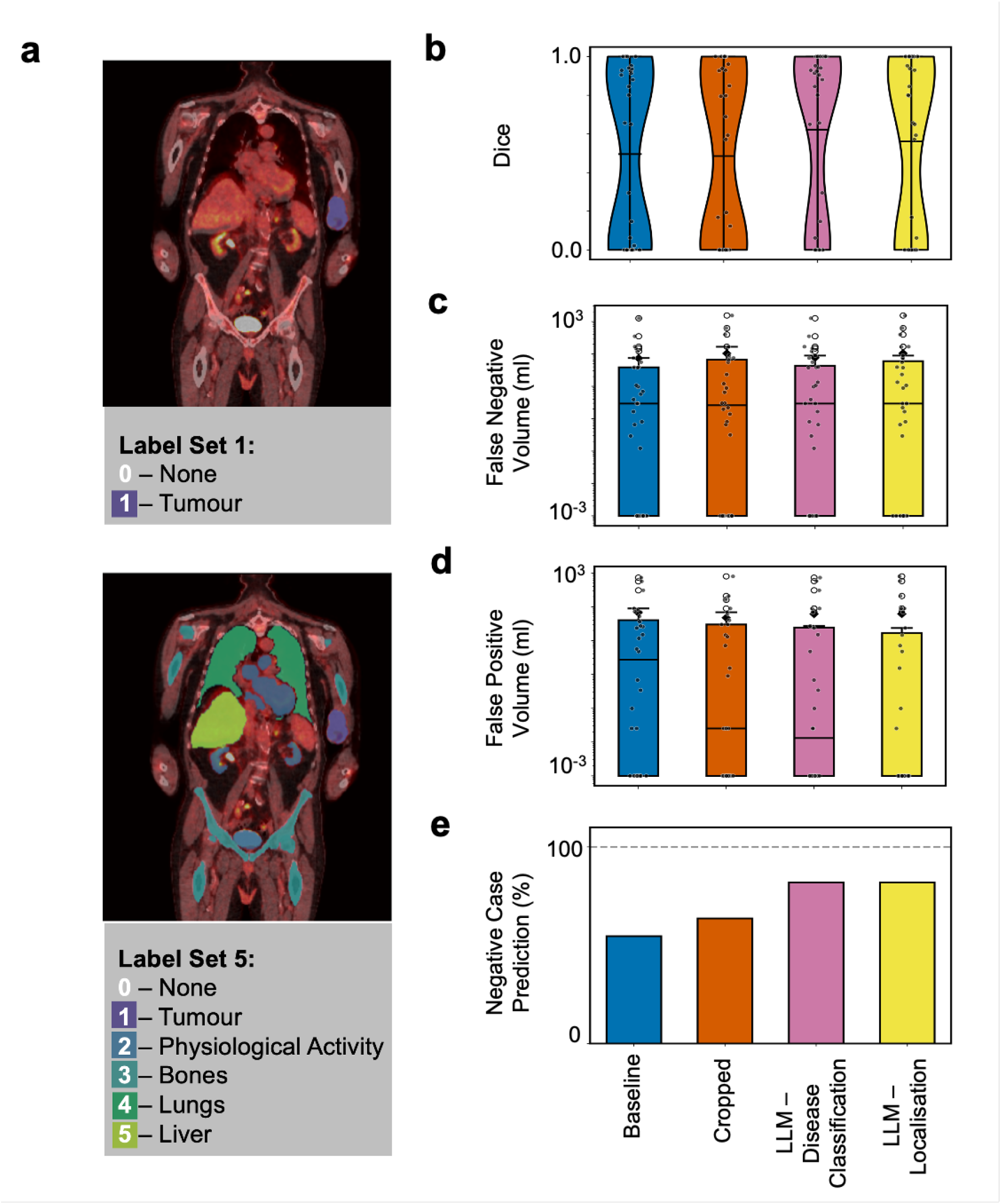
Overall Framework and Baseline Model Results. **a:** Example fused ^18^F-FDG PET/CT patient image with two different label sets used for training. Label set 1 was used in cropped models, with label set 2 used in models applied to the whole image. **b:** Violin plot of Dice coefficient across the test data for the different models/frameworks. **c:** Boxplot of the false negative volume across the different models/frameworks. **d:** Boxplot of the false positive volume across the different models/frameworks. **e:** Bar chart of the negative case prediction rate across the different models/frameworks.

The small number of cases the localisation framework is applied to arises from the confidence threshold required for this LLM task to achieve sufficient accuracy and thus high rejection rate (confidence threshold: 0.4, rejection rate: 0.76). Further research is required in the use of LLMs for classification and interpretation of complex unseen radiology reports. It is clear that medical benchmark datasets rarely represent this data adequately, leading to a gap in the field.

Using a small dataset with wide-ranging tumour localisation is a fundamental limitation. This work sought to explore what could be achieved with agentic framework methods in a limited dataset for a rare heterogeneous cancer. In particular, for the localisation task understanding the results is very challenging as the method was applied to such a small number of cases (N=8) in the test set. A larger dataset would enable exploration of higher levels of complexity at various stages throughout the framework. Withstanding additional complexity, this work demonstrates the potential of a relatively simple agentic framework, with logical clinical decision making built in, to improve results and step closer to acceptable performance for research implementation.

## 5 Conclusion

Incorporating radiology report data via a simple agentic framework improved automated segmentation performance in a sarcoma PET/CT dataset. Retrospective standard of care datasets contain radiology reports as standard and utilising this data could aid the use of imaging data for discovery research.

## Data Availability

The data that support the findings of this study are not openly available due to reasons of sensitivity and are available from the corresponding author upon reasonable request.

## References

[1] F. Isensee, P. F. Jaeger, S. A. Kohl, J. Petersen, and K. H. Maier-Hein, “nnU-Net: a self-configuring method for deep learning-based biomedical image segmentation,” Nature Methods, vol. 18, no. 2, pp. 203–211, Feb. 2021, issn: 15487105. doi: 10.1038/s41592-020-01008-z.

[2] S. Gatidis et al., “Results from the autoPET challenge on fully automated lesion segmentation in oncologic PET/CT imaging,” Nature Machine Intelligence, vol. 6, no. 11, pp. 1396–1405, Nov. 2024, issn: 25225839. doi: 10.1038/S42256-024-00912-9;TECHMETA. [Online]. Available: https://www.nature.com/articles/s42256-024-00912-9.

[3] N. Sourlos et al., “Recommendations for the creation of benchmark datasets for reproducible artificial intelligence in radiology,” Insights into Imaging, vol. 15, no. 1, pp. 1–12, Dec. 2024, issn: 18694101. doi: 10.1186/S13244-024-01833-2/TABLES/1. [Online]. Available: https://insightsimaging.springeropen.com/articles/10.1186/s13244-024-01833-2.

[4] T. Panch, T. J. Pollard, H. Mattie, E. Lindemer, P. A. Keane, and L. A. Celi, ““Yes, but will it work for my patients?” Driving clinically relevant research with benchmark datasets,” npj Digital Medicine, vol. 3, no. 1, pp. 1–4, Dec. 2020, issn: 23986352. doi: 10.1038/S41746-020-0295-6;SUBJMETA. [Online]. Available: https://www.nature.com/articles/s41746-020-0295-6.

[5] J. Whelan et al., “Incidence and survival of malignant bone sarcomas in England 1979-2007,” International journal of cancer, vol. 131, no. 4, Aug. 2012, issn: 1097-0215. doi: 10.1002/IJC.26426. [Online]. Available: https://pubmed.ncbi.nlm.nih.gov/21913189/.

[6] A. Bacon et al., “Incidence and survival of soft tissue sarcoma in England between 2013 and 2017, an analysis from the National Cancer Registration and Analysis Service,” International journal of cancer, vol. 152, no. 9, pp. 1789–1803, May 2023, issn: 1097-0215. doi: 10.1002/IJC.34409. [Online]. Available: https://pubmed.ncbi.nlm.nih.gov/36541754/.

[7] R. De Angelis et al., “The impact of radiomics in the management of soft tissue sarcoma,” Discover. Oncology, vol. 15, no. 1, p. 62, Dec. 2024, issn: 27306011. doi: 10.1007/S12672-024-00908-2. [Online]. Available: https://pmc.ncbi.nlm.nih.gov/articles/PMC10914656/.

[8] S. H. Kim et al., “Benchmarking the diagnostic performance of open source llms in 1933 eurorad case reports,” npj Digital Medicine, vol. 8, pp. 1–9, 1 Dec. 2025, issn: 23986352. doi: 10.1038/S41746-025-01488-3;SUBJMETA. [Online]. Available: https://www.nature.com/articles/s41746-025-01488-3.

[9] B. L. Guellec et al., “Performance of an Open-Source Large Language Model in Extracting Information from Free-Text Radiology Reports,” https://doi.org/10.1148/ryai.230364, vol. 6, no. 4, May 2024, issn: 26386100. doi: 10.1148/RYAI.230364. [Online]. Available: /doi/pdf/10.1148/ryai.230364.

[10] Q. Zeng et al., “Harnessing Text Insights with Visual Alignment for Medical Image Segmentation,” IEEE transactions on medical imaging, vol. PP, 2025, issn: 1558-254X. doi: 10.1109/TMI.2025.3601359. [Online]. Available: https://pubmed.ncbi.nlm.nih.gov/40839504/.

[11] Y. Sun et al., “Visual-language foundation models in medical imaging: A systematic review and meta-analysis of diagnostic and analytical applications,” Computer Methods and Programs in Biomedicine, vol. 268, p. 108 870, Aug. 2025, issn: 0169-2607. doi: 10.1016/J.CMPB.2025.108870. [Online]. Available: https://www.sciencedirect.com/science/article/pii/S0169260725002871.

[12] H. Lin, C. Xu, and J. Qin, “Taming Vision-Language Models for Medical Image Analysis: A Comprehensive Review,” Jun. 2025. [Online]. Available: https://arxiv.org/pdf/2506.18378.

[13] P. R. A. S. Bassi et al., “Learning segmentation from radiology reports,” in proceedings of Medical Image Computing and Computer Assisted Intervention – MICCAI 2025, vol. LNCS 15964, Springer Nature Switzerland, Sep. 2025.

[14] N. Dietrich, “Agentic AI in radiology: Emerging Potential and Unresolved Challenges,” The British journal of radiology, Jul. 2025, issn: 1748-880X. doi: 10.1093/BJR/TQAF173. [Online]. Available: https://pubmed.ncbi.nlm.nih.gov/40705666/.

[15] S. Wind et al., “Agentic large language models improve retrieval-based radiology question answering,” Lisa Adams, vol. 1, no. 2, Aug. 2025. [Online]. Available: https://arxiv.org/pdf/2508.00743.

[16] W. Chen et al., “RadFabric: Agentic AI System with Reasoning Capability for Radiology,” Jun. 2025. [Online]. Available: https://arxiv.org/pdf/2506.14142.

[17] G. K. Murugesan et al., “Improving Lesion Segmentation in FDG-18 Whole-Body PET/CT scans using Multilabel approach: AutoPET II challenge,” Nov. 2023. [Online]. Available: https://arxiv.org/pdf/2311.01574.

[18] A. Grattafiori et al., “The Llama 3 Herd of Models,” Jul. 2024. [Online]. Available: https://A.arxiv.org/pdf/2407.21783.

[19] A. Q. Jiang et al., “Mistral 7B,” Oct. 2023. [Online]. Available: https://arxiv.org/pdf/2310.06825.

[20] A. Yang et al., “Qwen2.5 Technical Report,” Dec. 2024. [Online]. Available: https://arxiv.org/pdf/2412.15115.

[21] M. S. Ankit Pal, Openbiollms: Advancing open-source large language models for healthcare and life sciences, https://huggingface.co/aaditya/OpenBioLLM-Llama3-70B, 2024.

[22] Y. Labrak, A. Bazoge, E. Morin, P.-A. Gourraud, M. Rouvier, and R. Dufour, Biomistral: A collection of open-source pretrained large language models for medical domains, 2024. 2402.10373[cs.CL].

[23] prithiv M Lmods, Qwen-umls-7b-instruct, https://huggingface.co/prithivMLmods/Qwen-UMLS-7B-Instruct, Hugging Face model card. License: CreativeML OpenRAIL-M. Accessed: 2025-07-26, 2025.

[24] J. Wasserthal et al., “TotalSegmentator: Robust Segmentation of 104 Anatomic Structures in CT Images,” Radiology: Artificial Intelligence, vol. 5, no. 5, e230024, Sep. 2023, issn: 26386100. doi: 10.1148/RYAI.230024. [Online]. Available: https://pmc.ncbi.nlm.nih.gov/articles/PMC10546353/.

[25] J. Wong et al., “Effects of interobserver and interdisciplinary segmentation variabilities on CT-based radiomics for pancreatic cancer,” Scientific Reports, vol. 11, no. 1, pp. 1–12, Dec. 2021, issn: 20452322. doi: 10.1038/S41598-021-95152-X;SUBJMETA. [Online]. Available: https://www.nature.com/articles/s41598-021-95152-x.

[26] C. Shyr et al., “A statistical framework for evaluating repeatability and reproducibility of large language models in diagnostic reasoning,” medRxiv, p. 2025.08.06.25333170, Aug. 2025, issn: 3067-2007. doi: 10.1101/2025.08.06.25333170. [Online]. Available: https://www.medrxiv.org/content/10.1101/2025.08.06.25333170v1 https://www.medrxiv.org/content/10.1101/2025.08.06.25333170v1.abstract.

[27] M. Omar, R. Agbareia, B. S. Glicksberg, G. N. Nadkarni, and E. Klang, “Benchmarking the Confidence of Large Language Models in Answering Clinical Questions: Cross-Sectional Evaluation Study,” JMIR Medical Informatics, vol. 13, e66917, May 2025. doi: 10.2196/66917. [Online]. Available: https://pmc.ncbi.nlm.nih.gov/articles/PMC12101789/.

[28] F. J. Dorfner et al., “Biomedical Large Languages Models Seem not to be Superior to Generalist Models on Unseen Medical Data,” Aug. 2024. [Online]. Available: https://arxiv.org/pdf/2408.13833v1.

[29] J. M. Nobel, K. van Geel, and S. G. Robben, “Structured reporting in radiology: a systematic review to explore its potential,” European Radiology, vol. 32, no. 4, p. 2837, Apr. 2021, issn: 14321084. doi: 10.1007/S00330-021-08327-5. [Online]. Available: https://pmc.ncbi.nlm.nih.gov/articles/PMC8921035/.

[30] V. Granata et al., “Structured Reporting of Rectal Cancer Staging and Restaging: A Consensus Proposal,” Cancers, vol. 13, no. 9, p. 2135, May 2021, issn: 20726694. doi: 10.3390/CANCERS13092135. [Online]. Available: https://pmc.ncbi.nlm.nih.gov/articles/PMC8125446/.

[31] S. A. Esfahani et al., “Standardized template for clinical reporting of PSMA PET/CT scans,” European Journal of Nuclear Medicine and Molecular Imaging, vol. 52, no. 1, p. 335, Dec. 2024, issn: 16197089. doi: 10.1007/S00259-024-06857-W. [Online]. Available: https://pmc.ncbi.nlm.nih.gov/articles/PMC11599343/.

