## Supplementary material for "Leveraging Large Language Models to Direct Automated PET/CT Tumour Segmentation in Retrospective Data: an Agentic Framework Method"

October 2025

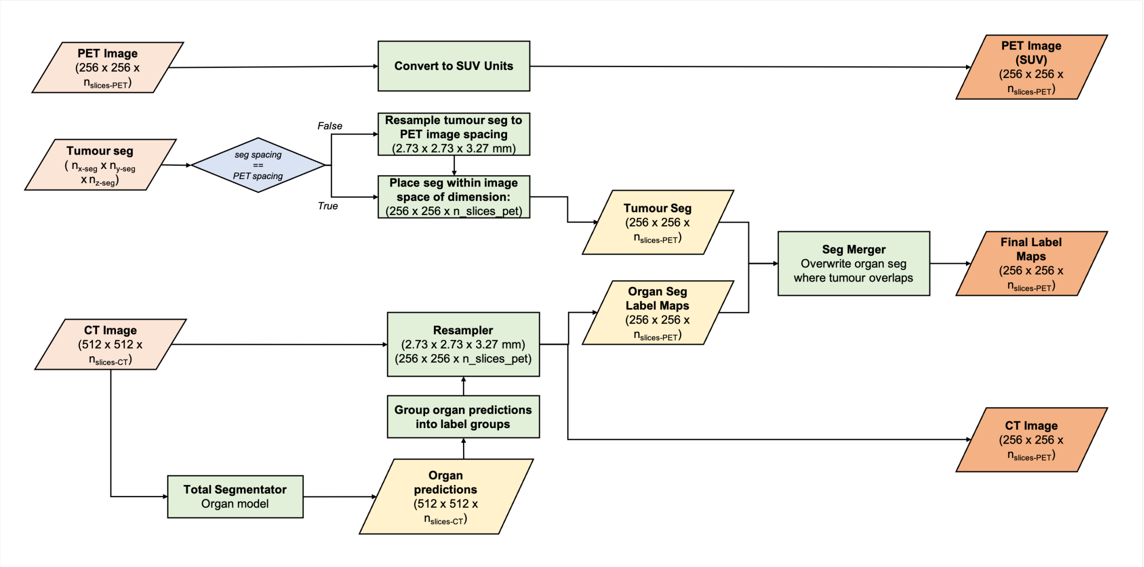


Figure 1: **Pre-processing for Image Segmentation**

The mapping for the different organ labels included in the four label groups is provided Table 1

| **Label Group** | **Total Segmentator Volumes Included** |
|---|---|
| Physiological | Heart, Urinary Bladder, Brain, Left Kidney, Right Kidney |
| Bone | Left Femur, Right Femur, Left Hip, Right Hip, Left Humerus, Right Humerus, Sternum |
| Lung | Left Lower Lobe, Right Lower Lobe, Right Middle Lobe, Left Upper Lobe, Right Upper Lobe |
| Liver | Liver |

Table 1: **Volumes included in grouped organ segmentations as generated from TotalSegmentator for nnUNet training.**

The organ labels were grouped into biologically relevant groups before being added to the training labels sequentially. The first group to be included in addition to tumour label data is the regions of expected high physiological uptake. The second group particularly relevant to sarcoma is bone, only large bones outputted by the Total Segmentator organ model are included, notably this includes lower leg and arm bones. The third group is lung relevant to sarcoma, as sarcoma frequently metastasises to the lung. The final inclusion of liver was chosen as this is a large volume in the abdomen that sarcoma very rarely metastasises to (no metastatic spread to liver present in training dataset).

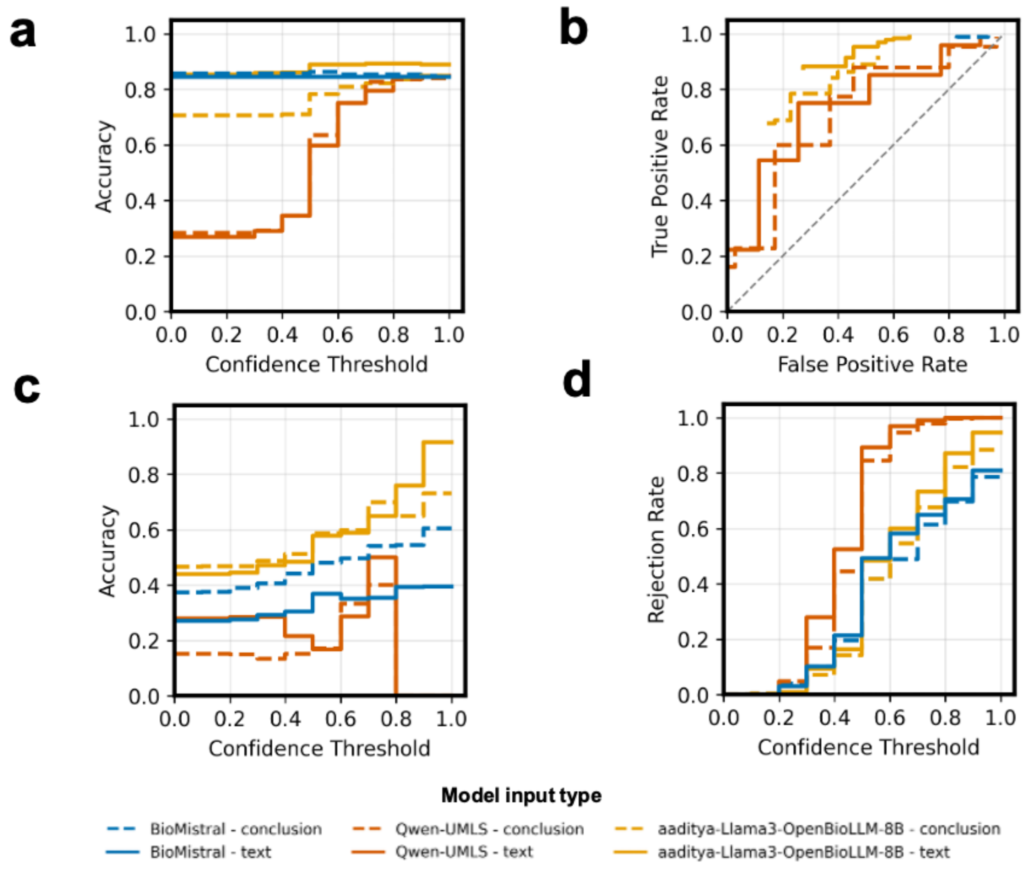


Figure 2: **LLM radiology report interpretation eesults for medically finetuned models.** **a:** Accuracy of the overall classification against confidence threshold. **b:** Receiver Operator Characteristic Curve for the overall process, the plot does not pass through (0,0) because the decision variable is not continuous. **c:** Accuracy of the localisation task as a function of confidence threshold. **d:** Rejection rate of the localisation task as a function of confidence threshold.

| Label Group | Total Segmentator Volumes Included |
|---|---|
| Head | Brain, Skull, Vertebrae C7, Vertebrae C6, Vertebrae C5 |
| Left Chest | Left Upper Lobe, Left Lower Lobe, Sternum, Vertebrae T1, Vertebrae T2, Vertebrae T3, Vertebrae T4, Vertebrae T5, Vertebrae T6, Vertebrae T7, Vertebrae T8, Vertebrae T9, Vertebrae T10, Vertebrae T11, Vertebrae T12 |
| Right Chest | Right Upper Lobe, Right Middle Lobe, Right Lower Lobe, Sternum, Vertebrae T1, Vertebrae T2, Vertebrae T3, Vertebrae T4, Vertebrae T5, Vertebrae T6, Vertebrae T7, Vertebrae T8, Vertebrae T9, Vertebrae T10, Vertebrae T11, Vertebrae T12 |
| Left Arm | Left Humerus |
| Right Arm | Right Humerus |
| Abdomen | Vertebrae T10, Vertebrae T11, Vertebrae T12, Vertebrae L1, Vertebrae L2, Vertebrae L3, Vertebrae L4, Vertebrae L5, Liver, Spleen, Left Kidney, Right Kidney, Sacrum |
| Pelvis | Sacrum, Left Hip, Right Hip |
| Left Leg | Left Femur, Left Hip |
| Right Leg | Right Femur, Right Hip |
| Lung | Left Upper Lobe, Left Lower Lobe, Right Upper Lobe, Right Middle Lobe, Right Lower Lobe |

Table 2: **Anatomical region mapping for Total Segmentator volumes.**
The organ segmentations from Total Segmentator are grouped into anatomically relevant regions for linking to the radiology report interpretation. Each region contains the relevant organs and structures within that anatomical area, with some structures (such as vertebrae and lung lobes) appearing in multiple regions where anatomically appropriate. The groupings facilitate region-specific analysis.

| Question ID | Question Text |
|---|---|
| 1.1 | Is there any evidence of FDG avid disease in the PET image? |
| 1.2 | Does this image show any evidence of cancerous disease? |
| 1.3 | Is there FDG-avid disease present in the scan? |
| 1.4 | Does this image show any FDG avid disease? |
| 1.5 | Is a tumour visible in the PET scan? |
| 1.6 | Are there any cancerous tumours present in the PET scan? |
| 1.7 | Does the PET image show any evidence of cancerous disease? |
| 1.8 | Is FDG uptake indicating disease visible in the scan? |
| 1.9 | Does the scan demonstrate FDG-avid disease? |
| 1.10 | Is abnormal FDG uptake consistent with disease seen in this scan? |
| 1.11 | Does the scan show FDG-avid disease? |
| 1.12 | Is FDG-avid disease present on PET/CT imaging? |

Table 3: **Disease classification task questions for LLM radiology report interpretation.**
All questions in this category are binary (Yes/No) queries about the presence of FDG-avid disease or cancerous tumours in PET/CT imaging.

| Question ID | Question Text |
|---|---|
| 2.1 | What is the location of any FDG avid disease present in the PET image? |
| 2.2 | Where is there FDG avid disease present in the PET/CT image? |
| 2.3 | In which body part is the FDG avid disease located in the PET image? |
| 2.4 | Where is the FDG avid disease located in the PET/CT image? |
| 2.5 | Which region of the body shows FDG-avid disease on PET/CT? |
| 2.6 | Where does the PET/CT show FDG uptake consistent with disease? |
| 2.7 | In which area is the disease located in the PET/CT image? |
| 2.8 | Which site demonstrates FDG-avid disease on PET/CT? |
| 2.9 | What is the location of FDG-avid disease in the PET/CT image? |
| 2.10 | Where is the FDG uptake located in the PET/CT image? |
| 2.11 | What is the location of any FDG uptake present in the PET image? |
| 2.12 | Where is there FDG uptake present in the PET/CT image? |

Table 4: **Localisation task questions for LLM radiology report interpretation.** All questions in this category ask about the anatomical location of disease, with standardized answer options covering nine body regions plus a negative option (Head, Left Chest, Right Chest, Left Arm, Right Arm, Abdomen, Pelvis, Left Leg, Right Leg, No disease)
